# An Accelerated PETALUTE MRI Sequence for *In Vivo* Quantification of Sodium Content in Human Articular Cartilage at 3T

**DOI:** 10.1101/2024.05.02.24305807

**Authors:** Cameron X. Villarreal, Xin Shen, Ahmad A. Alhulail, Nicholas M. Buffo, Xiaopeng Zhou, Evan Pogue, Ali Caglar Özen, Mark Chiew, Stephen Sawiak, Uzay Emir, Deva D. Chan

## Abstract

In this work, we demonstrate the sodium magnetic resonance imaging (MRI) capabilities of a three-dimensional (3D) dual-echo ultrashort echo time (UTE) sequence with a novel rosette petal trajectory (PETALUTE), in comparison to the 3D density-adapted (DA) radial spokes UTE sequence. We scanned five healthy subjects using a 3D dual-echo PETALUTE acquisition and two comparable implementations of 3D DA-radial spokes acquisitions, one matching the number of k-space projections (Radial – Matched Spokes) and the other matching the total number of samples (Radial – Matched Samples) acquired in k-space. The PETALUTE acquisition enabled equivalent sodium quantification in articular cartilage volumes of interest (168.8 ± 29.9 mM) to those derived from the 3D radial acquisitions (171.62 ± 28.7 mM and 149.8 ± 22.2 mM, respectively). We achieved a 41% shorter scan time of 2:06 for 3D PETALUTE, compared to 3:36 for 3D radial acquisitions. We also evaluated the feasibility of further acceleration of the PETALUTE sequence through retrospective compressed sensing with 2× and 4× acceleration of the first echo and showed structural similarity of 0.89 ± 0.03 and 0.87 ± 0.03 when compared to non-retrospectively accelerated reconstruction. Together, these results demonstrate improved scan time with equivalent performance of the PETALUTE sequence compared to the 3D DA-radial sequence for sodium MRI of articular cartilage.

## INTRODUCTION

Loss of glycosaminoglycan (GAG) from articular cartilage is an early hallmark of osteoarthritis [1–3]. GAGs, which are negatively charged, result in high cartilage fixed charge density, which is balanced by cations in the interstitial fluid. Sodium (^23^Na) magnetic resonance imaging (MRI) signal can be used as a direct measure of GAG content [4, 5], through its direct relationship maintaining the charge balance against the negative tissue fixed charge density [6]. Therefore, ^23^Na MRI could provide valuable diagnostic information during the early stages of osteoarthritis progression before the onset of pain or radiographic joint space narrowing, as quantified by the Kellgren-Lawrence grading scale [7]. Despite the potential of ^23^Na MRI as a powerful diagnostic tool for early osteoarthritis, its practical use in the clinic is hampered by long scan times, poor spatial resolution, and low inherent sodium content, which typically prompts the need for higher magnetic field strength [8, 9]. Additionally the physical properties of sodium (i.e., 3/2 spin quadrupolar interaction, fast T_2_ relaxation, gyromagnetic ratio [10]) and its lower abundance lead to reduced sensitivity and correspondingly low signal-to-noise ratio (SNR) [11]), make it necessary to use an ultrashort echo time (UTE) sequence. The low thickness of articular cartilage in healthy adult humans [12] also presents a challenge for sodium MRI, since high spatial resolution, and therefore longer scan times, is preferred for both avoiding partial volume effects and measuring spatially varying sodium signal in cartilage.

Advancements in UTE sequences have enabled better imaging and quantification of sodium in vivo. UTE sequences enable adequate coverage of k-space before substantial decay of the signal (T_2_* of 13.2 ms [13]), although the trajectories with which they traverse k-space differ. Some common 3D trajectories include cones, spiral, and density-adapted (DA) radial spokes [9, 13, 14]. Currently, the 3D radial acquisition is often employed for sodium imaging in cartilage due to its ability to rapidly acquire signal at high resolution with relatively fast echo time. This results in a high-quality sodium signal without sacrificing acquisition time. The 3D radial acquisition has also been used for in vivo quantifying sodium concentration in cartilage at 3T and 7T. Recently, Shen, *et. al.*, developed a novel 3D rosette petal trajectory UTE (PETALUTE) sequence that features a curvilinear, petal-like trajectory, in contrast to the linear trajectory of a radial spoke [15]. Using proton MRI of the brain, they showed that 3D dual-echo PETALUTE trajectory achieved comparable coverage of k-space and signal quality as 3D radial at a shorter total acquisition time [15].

In this work, our objective was to evaluate the PETALUTE sequence for *in vivo* sodium MRI of the human knee and sodium content quantification in articular cartilage. Toward the goal of addressing several limitations to sodium MRI, we demonstrate that the 3D dual-echo rosette acquisition reduces scan time while preserving signal quality when compared to the 3D DA-radial spokes acquisition. Furthermore, we demonstrate the ability to further accelerate the PETALUTE acquisition via compressed sensing without significant reduction in signal quality.

## METHODS

### 3D Dual-Echo UTE MRI with novel Rosette k-Space Trajectory (PETALUTE)

We implemented a novel rosette trajectory for a 3D UTE acquisition that traverses k-space with multiple crossings of the k-space origin (**Figure 1**) [15]. Briefly, the petal-like sampling enabled a dual echo acquisition, with petal trajectory passing through k-space center at the start and end of each repetition. Images were reconstructed from the first and second half of each petal. 210 samples per petal and 18050 petals are acquired with an acceleration factor of 2. Due to the efficient sampling of the novel rosette k-space pattern, only 80% of the required k-space (a total of 31600 samples) can be considered full-k-space acquisition [15].

**Figure 1:**
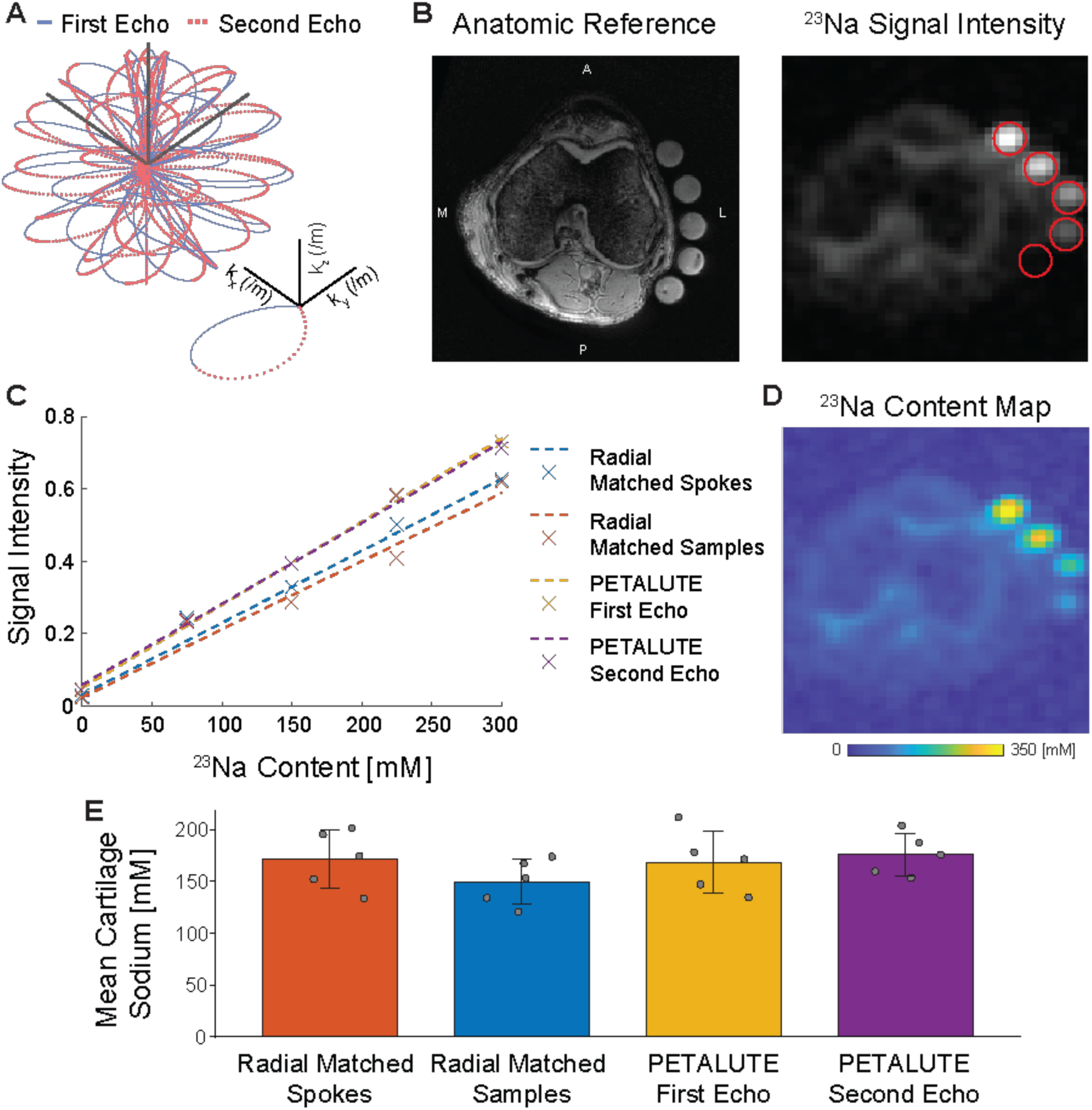
Dual-echo PETALUTE was compared to Radial UTE for 3D acquisition of sodium quantification in articular cartilage. (A) 3D dual-echo PETALUTE is a novel k-space trajectory that enables a more efficient sampling of the edges of k-space. The outward (solid blue) and inward (dashed red) petal-like trajectories enable acquisition of dual-echo images. (B) Cartilage regions of interest were manually segmented using on the anatomical reference for each subject. Subjects and sodium concentration standards (red circles) were scanned with 3 different sodium MRI acquisitions: 3D dual-echo PETALUTE (representative axial slice shown), Radial – Matched Samples, and Radial – Matched Spokes. (C) A linear curve fit was used to calibrate sodium signal intensity. (D) Signal intensity was then converted to sodium concentration maps. (E) Sodium concentrations were averaged over the cartilage ROIs for each subject and UTE acquisition, with mean ± standard deviation concentrations also shown.

### Subject Selection and Scan Parameters

Under Institutional Review Board approval, we scanned the right knees of five healthy volunteers (29.6 ± 13.4 years old, 1 female) in a Siemens MAGNETOM Prisma 3-Tesla MRI scanner (Siemens Healthineers, Germany). We positioned each subject supine and foot first and used a goniometer to flex the knee at 15° before adding foam padding and securing the dedicated knee coil. With the joint midline (identified via palpation) aligned to isocenter, ^1^H anatomical reference scans were acquired using the integrated body coil to mitigate registration error between proton and sodium images. All volunteers self-reported no history of joint trauma or disease and showed no indications of musculoskeletal disease on anatomical scans.

Using a frequency-tuned, mono-resonant ^23^Na transmit-receive knee volume coil (32.6 MHz, Stark-Contrast, Erlangen, Germany), we compared the 3D dual-echo rosette sequence [15] to two implementations of the 3D DA-radial spokes sequence (**Table 1**). We adjusted the radial sequence to either match the same number of points sampled in k-space (“Radial – Matched Samples”) or the same number of projections into k-space (“Radial – Matched Spokes”). Comparisons to DA-radial acquisitions were performed against one of two images acquired with the 3D rosette dual-echo sequence (“Rosette – First Echo”, “Rosette – Second Echo”). The lowest possible repetition time (TR) and echo time (TE) were chosen for each sequence.

**Table 1.** Acquisition parameters for each of the UTE sequences compared in the study.

| UTE Sequence Parameters | Dual-echo Rosette (PETALUTE) | Radial – Matched Samples | Radial – Matched Spokes |
| --- | --- | --- | --- |
| Number of averages (NA) | 3 | 3 | 3 |
| Repetition time (TR) | 7 ms | 12 ms | 12 ms |
| Number of PETAL/Spokes | 18050 | 18050 | 18050 |
| Number of Points per petals/spokes | 218 | 256 | 384 |
| RF Pulse Duration | 100 μs | 500 μs | 500 μs |
| ADC Bandwidth | 100 kHz | 25 kHz | 38 kHz |
| Echo time (TE) | 90 μs, 2270 μs | 300 μs | 300 μs |
| 3D Field of view (FOV) | 360 mm | 320 mm | 320 mm |
| Ernst angle | 44° | 55° | 55° |
| Acquisition time (mm:ss) | 2:06 | 3:36 | 3:36 |

### Sodium concentration standards

We prepared sodium phantoms using NaCl (0, 75, 150, 225, 300 mM) and 10% agarose w/v in distilled water in 15 mL conical tubes to generate a series of concentration standards that encompass the expected physiological range of sodium which has been measured up to ∼280 mM within healthy human femoral cartilage [6, 16]. We secured the phantoms to the lateral aspect of each subject’s knee for scanning, and they were included in the field of view during all scans.

### Image Reconstruction and sequence comparisons

We reconstructed images via regular regridding applying a density-compensated adjoint nonuniform fast Fourier transform using the BART toolbox [17] in MATLAB (MathWorks, USA). We used a non-uniform fast Fourier transform (NUFFT) [18] to calculate the forward-encoding transform of the acquired k-space information acquired with the rosette UTE sequence. [19]. For PETALUTE acquisitions a compressed sensing approach was used for image reconstruction using total generalized variation as the sparsifying penalty [18, 20]. For both PETALUTE (acceleration factor of 2) and DA-radial we applied a Hanning filter for regular regridding, density compensated adjoint NUFFT. The resulting reconstructions resulted in a nominal resolution of 2.81 mm (isotropic). We determined the signal to noise ratio by dividing the average signal intensity value in cartilage regions of interest by the mean signal intensity of the phantom with 0 mM sodium concentration, which should be associated with noise in a sodium MRI acquisition.

### Sodium Quantification

To calibrate sodium concentration to acquired signal intensity, we identified the regions occupied by our sodium concentration standards, quantified the mean intensity value within each using five representative slices, and performed a linear curve fit to obtain a standard curve for each acquisition. Using this linear relationship, we performed voxel-wise sodium signal-to-concentration conversion. We manually segmented cartilage using anatomical reference scans and used these regions of interest to quantify the sodium concentration within cartilage (**Figure 1**).

### Retrospective Compressed Sensing

We applied compressed sensing of rosette acquisitions retrospectively using acceleration factors of 2× and 4× by pseudo-random under-sampling of k-space. We pseudo-randomly utilized one-half and one-quarter of the petals to reconstruct dual-echo images using the same reconstruction pipeline described previously. These simulated data sets were compared to dual-echo images with no acceleration factor using a structural similarity (SSIM) coefficient (*ssim* function, MATLAB, MathWorks). For each subject, we compared a cuboid volume of interest that contained the knee and determined the mean SSIM coefficient across all subjects.

### Statistical Analysis

All results are reported as mean ± standard deviation. We compared mean sodium concentrations using a Kruskal-Wallis test and evaluated the agreement between cartilage sodium concentration using a Bland-Altman test, comparing pixel-wise within each subject and as averaged values for cartilage in each subject. We used the PETALUTE – First Echo as our reference (all difference values were obtained by subtracting the sodium signal of each acquisition from the PETALUTE – First Echo values). The limits of agreement represent a 95% confidence interval centered on the mean difference for each comparison. Statistical analyses were performed in Rstudio [21], a development environment for R [22], using native functions and the *blandr* package [23].

## RESULTS

### Sequence Comparisons

Dual-echo PETALUTE and DA-radial acquisitions (matched by samples and by number of spokes) were used for sodium MRI of the knee in 5 human volunteers (**Figure 2**). The Radial Matched-Samples acquisition produced a ringing artifact in all subjects that was not observed in either of reconstruction for the dual echo PETALUTE sequence nor Radial Matched-Spokes acquisition. Calibration using sodium standards enabled calculation of sodium content maps (**Figure 3**). For the same number of averages PETALUTE allowed us to achieve a 41% reduction in total scan time. The SNR in knee cartilage was determined to be 9.2 ± 3.2 and 8.2 ± 2.9 for the first echo of the PETALUTE and Radial-Matched Spokes, respectively (p = 0.62).

**Figure 2:**
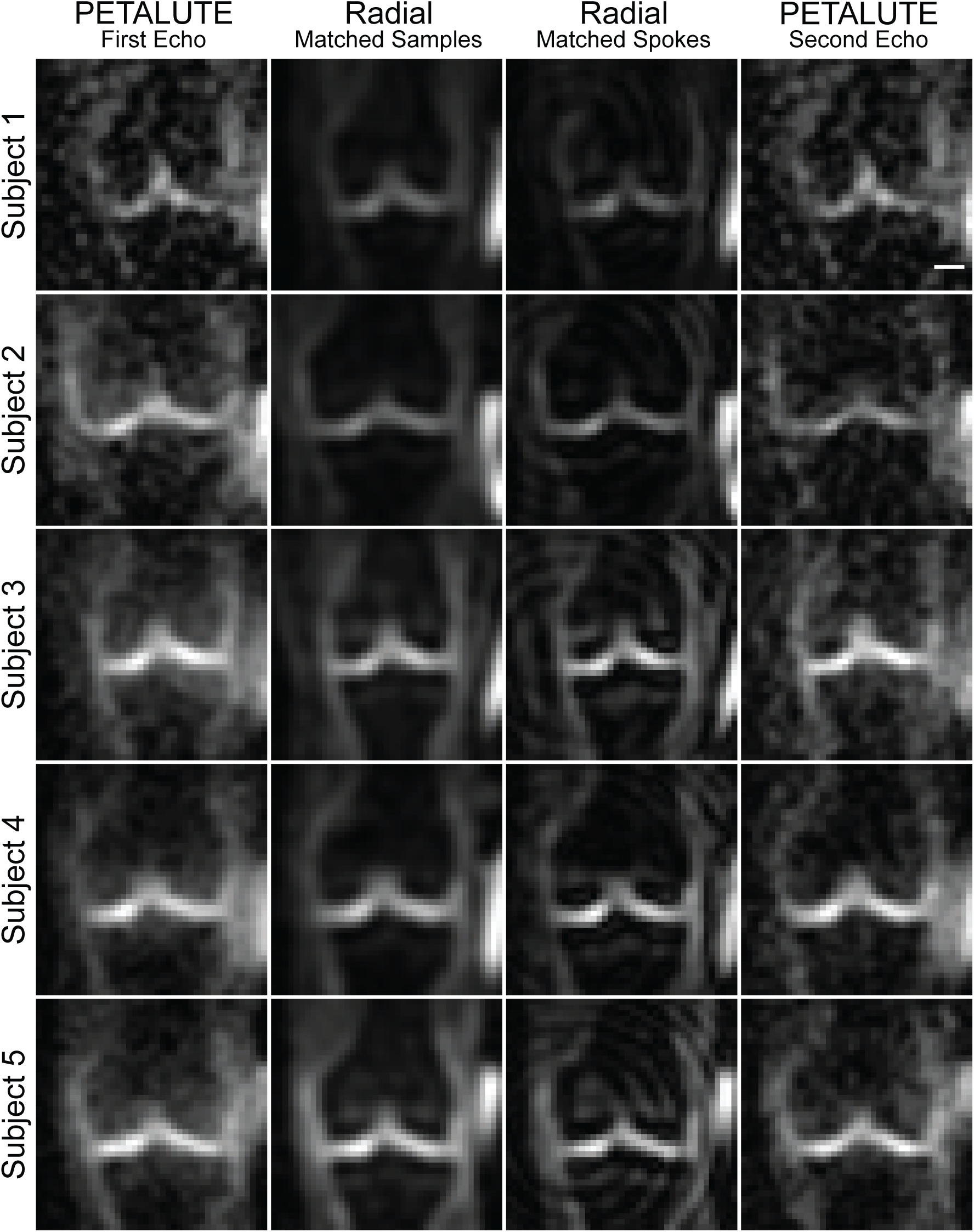
^23^Na MRI of the knee was implemented using PETALUTE and DA-Radial UTE. Cuboid cropped volumes were centered on the joint midline and include the distal femur and proximal tibia. Coronal slices centered on the joint space are shown for each subject and UTE acquisition. Scale bar = 1.5 cm.

**Figure 3:**
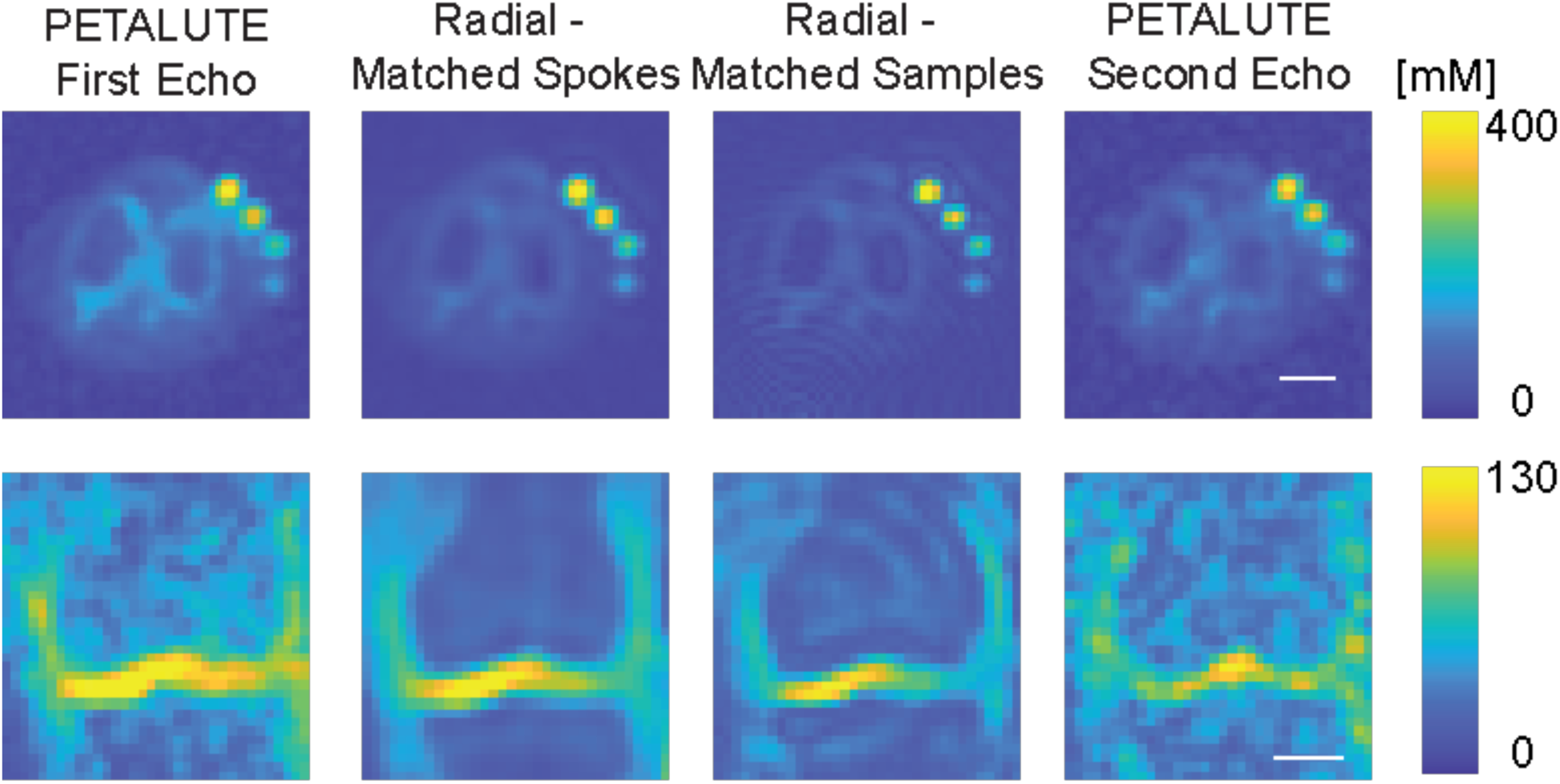
Sodium maps derived from dual-echo PETALUTE and Radial UTE were compared. Representative sodium maps are shown for a single subject for each of the UTE acquisitions. The axial images are cropped to 14.05 cm × 14.05 cm so that all phantoms and the knee are in view. Separately, a coronal slice, cropped to 11.2 cm × 11.2 cm of the same subject is shown, focusing on visualization of the tibial and femoral cartilage for medial and lateral compartments, allowing for a color range that is not dominated by the sodium phantom signal. Scale bar = 2.5 cm.

### Sodium Quantification

We calculated mean sodium values within cartilage of 173.0 ± 27.4 mM for PETALUTE– First Echo, 180.2 ± 34.3 mM for Radial – Matched Spokes, 146.7 ± 21.6 mM, and for Radial – Matched Samples (**Figure 1**). Sodium concentration maps derived from PETALUTE acquisition from which we subtracted the Radial UTE acquisitions to determine spatially dependent differences in the acquisitions (**Figure 4A**). The mean differences in measured cartilage sodium content between PETALUTE– First Echo and Radial – Matched Samples and Radial – Matched Spokes was 26.34 ± 15.67 mM and −7.146 ± 29.8 mM, respectively, and were not statistically significant (p = 0.199 and p = 0.668, respectively). The difference in measured sodium concentration between the first and second echo of PETALUTE was 12.99 ± 16.65 mM, which was not statistically significant (p = 0.656). Bland Altman mean-difference test showed that the limits of agreement for sodium concentration between each of the sequences bounded zero for pixel-wise comparisons within each subject (**Supplemental Figure 1**) and for mean cartilage sodium content across all subjects (**Figure 4B**).

**Figure 4:**
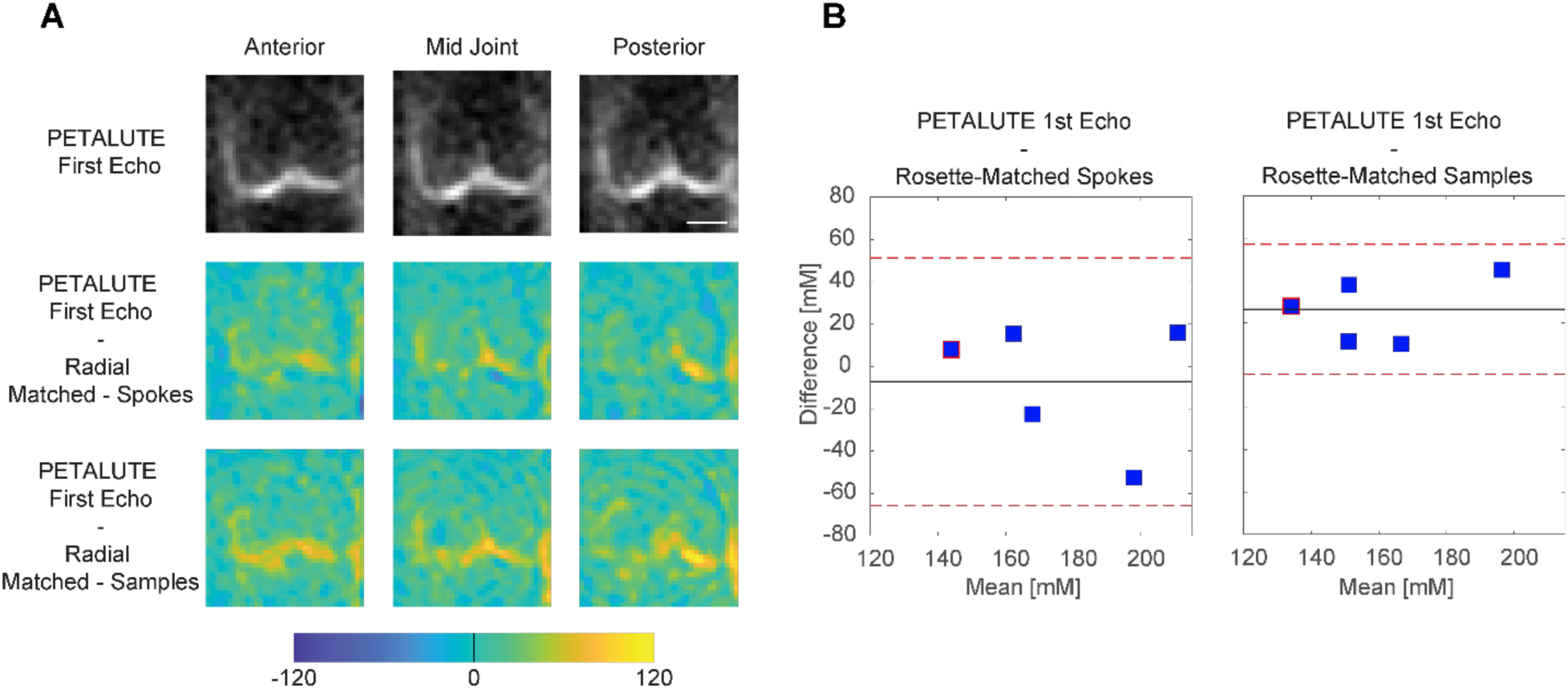
Sodium quantification with the PETALUTE and radial acquisitions shows strong agreement. The PETALUTE and radial acquisitions perform similarly for sodium content quantification. (A) Differences in sodium measurements were calculated by subtracting the sodium maps, derived from a radial acquisition, from the sodium map derived from the PETALUTE – First Echo acquisition. Three images from a representative subject are shown, the first and last slices (anteriorly and posteriorly) in which femoral cartilage is present and the center of the joint. The reference PETALUTE – First Echo acquisition is shown in the first row of images. (B) Bland-Altman test was performed to determine the limits of agreement between each combination of sequences, using PETALUTE – First Echo as the reference. Limits of agreement are denoted by red dashed horizontal lines, while the mean difference is denoted by a solid black line. For each subject, the mean and difference in sodium concentration in knee cartilage are plotted for the comparisons between PETALUTE – First Echo and Radial – Matched Spokes Samples and Radial – Matched Samples. The data points for the representative subject are indicated in red on these plots.

### Retrospective Compressed Sensing

We retrospectively evaluated imaging acceleration by down-sampling unaccelerated acquisitions prior to compressed sensing reconstruction. Using unaccelerated PETULATE – First Echo as the reference, SSIM indices were found to be 0.93 ± 0.02 and 0.89 ± 0.03 for 2× and 4× acceleration factors, respectively. For PETULATE – Second Echo, SSIM indices were 0.90 ± 0.02 and 0.87 ± 0.03, respectively (**Figure 5**).

**Figure 5:**
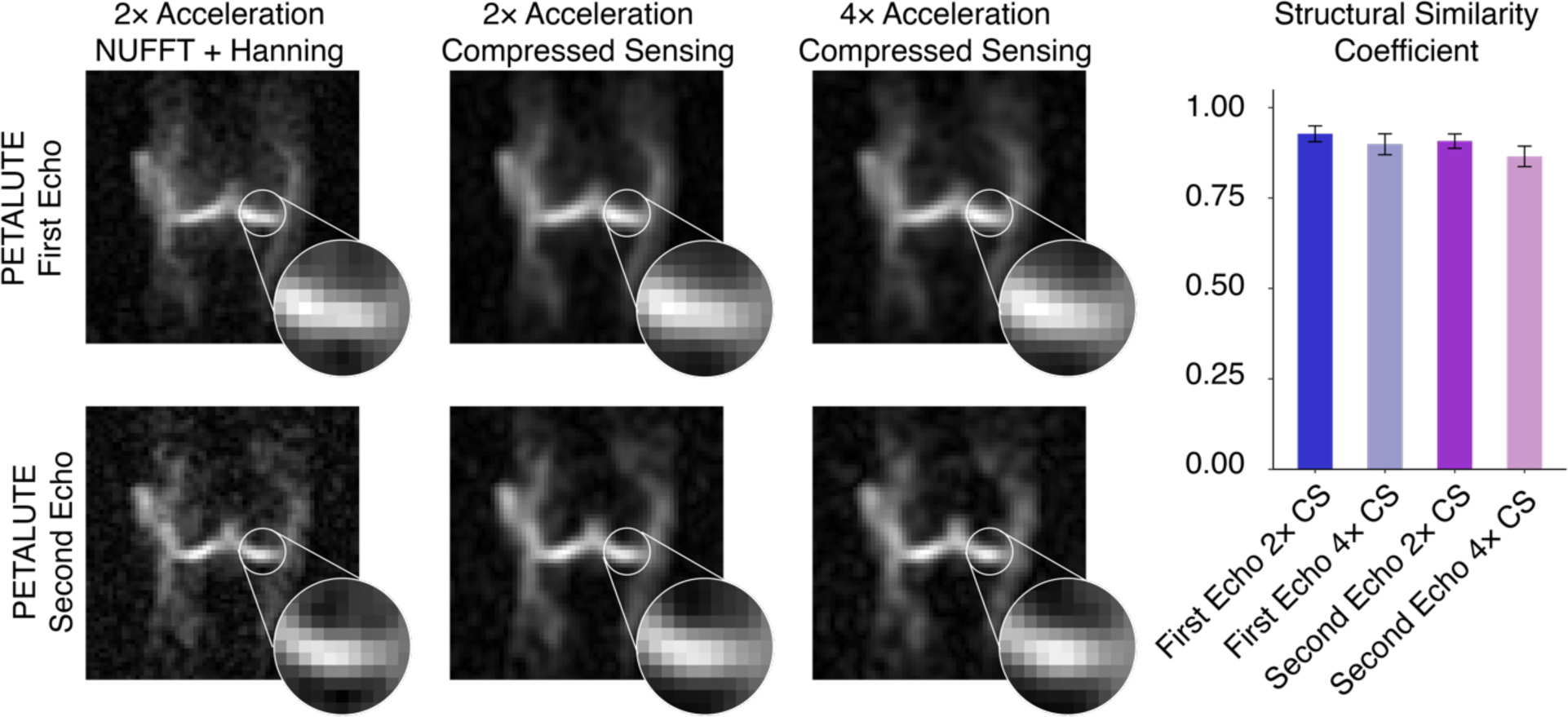
Retrospective compressed sensing demonstrates the capability for at least four-times acceleration while preserving image quality of dual-echo PETALUTE acquisitions. (A) Retrospective down-sampling of unaccelerated PETALUTE acquisitions were used to evaluate 2× and 4× acceleration via compressed sensing. (B) Structural similarity indices were calculated for each subject and both echoes showed high agreement with unaccelerated images. Qualitatively, boundaries between regions of high and low sodium signal intensity retain their similarity even at higher levels of acceleration.

## DISCUSSION

In this study, we demonstrate for the first time a 3D dual-echo novel rosette acquisition, PETALUTE [15], for sodium UTE MRI of the human knee. Using 3D PETULATE, we achieved rapid acquisition of sodium signal with high agreement in sodium quantification in articular cartilage with the 3D radial acquisition, with a 41% improvement in total acquisition time (**Table 1**). Additionally, acceleration of 2× and 4× via a simulated compressed sensing acquisition showed high structural similarity to the non-accelerated, full k-space rosette acquisition. Our results demonstrate that the novel 3D dual-echo rosette sequence [15] can rapidly acquire sodium signal with a high degree of agreement with the radial spokes acquisition and at faster total scan time.

To demonstrate the application of PETULATE for sodium MRI of the knee, we compared the 3D dual-echo PETALUTE sequence to 3D DA-radial spokes sequence. The radial spokes acquisition is commonly used in musculoskeletal imaging [8], and has been well documented for *in vivo* sodium imaging, particularly in imaging knee cartilage as in this work. For example, Madelin, *et. al.*, performed 3D radial MRI at 7T in human knee cartilage [24] which enabled sodium quantification in vivo, but even with 7T field strength, scan times exceeded 16:50. The 3D radial acquisition has also been used at 3T to quantify sodium content in the knee [25], but a total acquisition time of 57:45 was required to achieve a 3-mm isometric voxel size with one average. Across all subjects, the Radial – Matched Samples acquisition displayed a pronounced ringing artifact that was not evident in either Radial – Matched Spokes or PETALUTE (**Figure 1**), an effect likely attributable to undersampling. On the other hand, the rosette trajectory of PETALUTE provides improved k-space coverage which enables undersampling without losing image quality [15, 26]. Because of this improved efficiency, PETALUTE enabled an appreciably shorter total scan time of 2:06 (min:sec) compared to 3:36 while not significantly affecting sodium concentration measurements (**Figure 1**).

We achieved comparable SNR despite the shorter total scan time, with a mean SNR in knee cartilage of 9.2 using the first echo of PETULATE and 8.2 for Radial – Matched Spokes. Other studies have reported SNR of 30 in knee cartilage for the 3D radial sequence at 3T with a total acquisition time of 20 minutes [27]. In another study, 3D cones achieved an SNR of 11.3 in patellar cartilage at 3T with a total scan time of 25:50 [13]. Although direct comparisons of SNR cannot be made without matching all image parameters on the same system in the same subjects, increased averaging with PETALUTE – potentially in combination with compressed sensing – could readily match the SNR of previously reported studies. For example, one study employed a 3D radial with an acquisition 17 minutes and required a 7T field strength with an SNR in patellar cartilage of 43.8 ± 7.5 [28]. Since SNR is directly proportional to field strength and the square root of the number of averages, PETALUTE using the parameters in this 3T study with 8 averages (matching total acquisition time of about 17 min) would be expected to exceed the SNR (61.1 compared to 43.8). Furthermore, we show that PETALUTE performs with high structural similarity even with acceleration by compressed sensing (**Figure 5**), enabling further reductions in total scan time.

Beyond total scan time and SNR, PETALUTE also performed as well as the 3D DA-radial UTE sequence for quantifying sodium signal in articular cartilage *in vivo*. Using PETALUTE acquisition, we found no statistical difference (p = 0.345) in the sodium concentration quantified from the Radial – Matched Spokes acquisition (**Figure 1**), a well-established method for *in vivo* cartilage sodium quantification [25]. A Bland-Altman test showed limits of agreement that bounded zero and indicated a small, statistically insignificant difference in mean sodium content between the PETALUTE and radial UTE acquisitions.

These findings illustrate the capabilities of the novel 3D dual-echo rosette sequence to match the performance of the 3D radial acquisition while improving scan time, addressing one of the critical barriers to clinical translation of sodium MRI. However, all sequences tested in our study resulted in estimated sodium concentrations below the expected *in vivo* sodium concentration of cartilage.

Other works, which reported similarly unexpectedly low signal in healthy tissue, attributed this effect to the high water content (∼75% total weight) of cartilage [27, 29]. Accordingly, a correction factor of 1/0.75 has been proposed for cartilage to account for this sodium signal attenuation [24]. The average sodium concentration would have fallen within the expected physiological range for all acquisitions if we had adjusted concentrations by the correction factor. However, it is important to note that as water content in the cartilage changes with the loss GAG during early osteoarthritis [30], a constant calibration factor may not be appropriate in diagnostic practice.

Interestingly, we noted qualitative differences in image quality and sodium concentration mapping between the PETALUTE and the radial spokes acquisitions, although these could be attributed to the sampling approach in the Radial – Matched Samples acquisition. This approach thesame number of points sampled in k-space compared to PETALUTE, but due to the trajectory of the radial spokes this resulted in sampling near the Nyquist frequency. Additionally, we noted a difference in the sodium signal distribution in articular cartilage, particularly in the outer regions of the knee, between PETALUTE and radial UTE. In the sodium content difference images (**Figure 4**), higher sodium content is measured at mid-joint with PETALUTE than Radial – Matched Spokes. Additionally, we observed that the interfaces between tissue (ie: cartilage to bone), and the medial regions of femoral cartilage, tend to be sites of greater magnitude of sodium signal difference (**Figure 4**). With the radial acquisitions, the sodium signal intensity appeared to attenuate with distance from the center of the knee. However, in healthy articular cartilage, PG content, and thus the fixed charge density and sodium distribution, is not expected to differ substantially by location in the joint [31]. In contrast, the PETALUTE acquisition showed similar sodium signal intensity in the inner and outer regions of the joint, suggesting no signal drop-off further from the center of the joint. Although we cannot validate these sodium maps against “ground truth” PG content in these *in vivo* studies, it may be presumed that PG content in cartilage does not vary substantially by location within cartilage of a healthy knee joint [31]. Thus, these qualitative comparisons suggest that PETALUTE may more accurately render sodium concentration throughout the knee, particularly in regions more removed from the center of the image volume. This could be due to the greater density of sampling of outer k-space [32] where finer details, sharp transitions, and other higher frequency information is located. We observed a similar relationship in the retrospective compressed sensing study. There is no qualitative difference in signal pattern at the cartilage margins in any of the retrospective accelerations compared to the unaccelerated scans (**Figure 5**). The sharp signal gradients at the cartilage margins are preserved, even under 4× acceleration, for both echoes, suggesting minimal loss of high-frequency information.

In conclusion, sodium MRI has potential to become a powerful early diagnostic tool in cartilage degeneration, but its clinical translation is hindered by several factors, including long scan times and poor spatial resolution. In this work, we evaluated PETALUTE, a novel 3D rosette k-space trajectory, for sodium MRI of articular cartilage and showed reduced scan time compared to comparable radial acquisitions with a high degree of agreement in sodium quantification. Using retrospective analyses, we also demonstrated potential for greater acceleration of PETALUTE for sodium MRI through compressed sensing. Sodium MRI with 3D dual-echo PETALUTE acquisition therefore addresses several of the current barriers for clinical translation of sodium quantification in healthy and degenerating articular cartilage.

## Data Availability

All data produced in the present study are available upon reasonable request to the authors.

https://purr.purdue.edu

## ACKNOWLEDGMENTS

We thank Armin M. Nagel for providing the radial sequence for comparison. Data acquisition was supported in part by NIH grant S10 OD012336. In addition, D.D.C. was supported in part by NSF grant 2149946, and C.X.V. was supported in part by NIH training grant T32 DK101001.

## COMPLIANCE WITH ETHICAL STANDARDS

All procedures performed in studies involving human participants were in accordance with the ethical standards of the institutional and/or national research committee and with the 1964 Helsinki declaration and its later amendments or comparable ethical standards. Informed consent was obtained from all individual participants in the study, including permission to report identifying information, including images, in this article.

