## Supplemental Figure 1 for "An Accelerated PETALUTE MRI Sequence for *In Vivo* Quantification of Sodium Content in Human Articular Cartilage at 3T"

### SUPPLEMENTAL INFORMATION

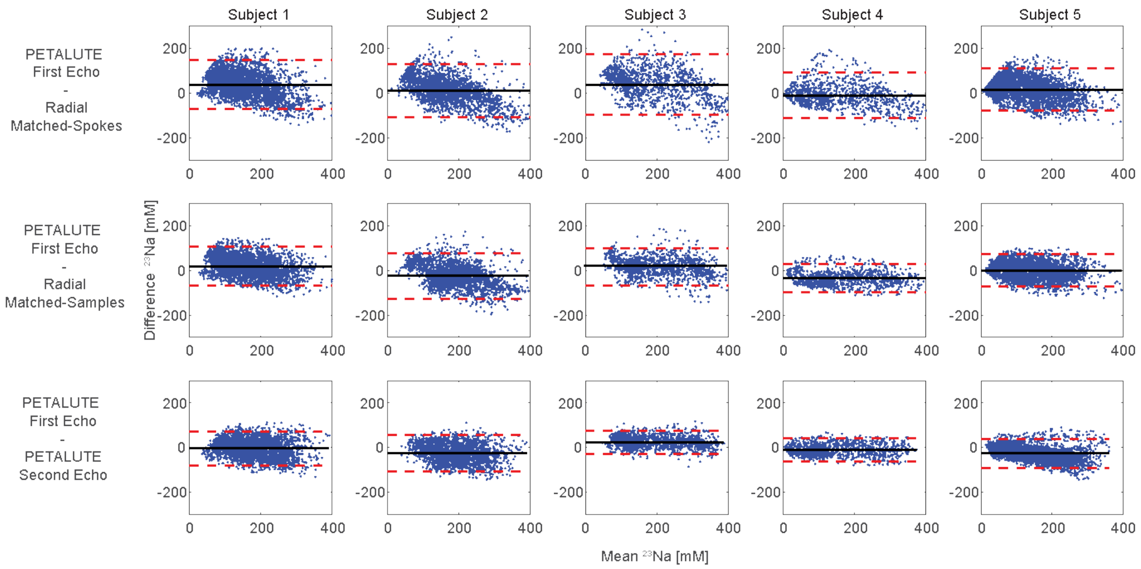


**Supplemental Figure 1: Bland Altman comparison indicates strong pixelwise agreement between PETALUTE and radial acquisitions.** The PETALUTE and the radial acquisitions perform similarly for sodium content quantification. Bland-Altman limits of agreement comparisons for each subject and sequence. Limits of agreement are denoted by red horizontal lines with confidence intervals for the limits of agreement denoted by dashed black lines. For each subject, the mean and difference in sodium concentration are plotted for the comparisons between Rosette – First Echo and Radial – Matched Samples (first row), Radial – Matched Spokes (second row), and Rosette – Second Echo (third row).
